# Psychometric validation of the Obstetric Quality of Recovery-10 scoring tool across the first month postpartum: a cross-sectional psychometric study

**DOI:** 10.64898/2026.07.08.26357380

**Authors:** E. Celetta, E. Lorthe, G. Cattani, M. Epiney, S. Grylka-Baeschlin, A. N. Mueller, J. Sormani, M. Suppan, I. N. Widmer, L. Gaucher, T. Desplanches

**Affiliations:** Geneva School of Health Sciences, HES-SO University of Applied Sciences and Arts Western Switzerland, Geneva, Switzerland; Research on Healthcare Performance (RESHAPE), Université Claude Bernard Lyon 1, INSERM U1290, Lyon, France; Université Paris Cité and Université Sorbonne Paris Nord, Inserm, INRAE, Centre for Research in Epidemiology and Statistics, F-75004 Paris, France; Division of Obstetrics, department of Paediatrics, Gynecology and Obstetrics, Faculty of Medicine, Geneva University Hospitals, Geneva, Switzerland; Research Institute of Midwifery and Reproductive Health, Zurich University of Applied Sciences, Winterthur, Switzerland; Midwifery Division, Bern University of Applied Sciences, Bern, Switzerland; Division of Anesthesiology, Department of Acute Medicine, Geneva University Hospitals, Geneva, Switzerland; Department of Anesthesiology, Pharmacology, Intensive Care and Emergency Medicine, Faculty of Medicine, University of Geneva, Geneva, Switzerland

**Keywords:** ObsQoR, Patient Reported Outcome Measures, Postpartum Period, Postpartum recovery, Psychometrics, Recovery of Function

## Abstract

**Background:** Postpartum recovery is a public health concern. The Obstetric Quality of Recovery-10 (ObsQoR-10) is a brief patient-reported outcome measure designed to assess early recovery after childbirth. Its validation is currently limited to the first three days postpartum. This study aimed to evaluate the psychometric properties of the ObsQoR-10 across the first 30 days postpartum.

**Methods:** We conducted a cross-sectional psychometric evaluation of the ObsQoR-10 using baseline data from a national Swiss multilingual cohort (French, German, Italian, and English). Women were recruited within the first week postpartum and completed the ObsQoR-10 and the EuroQol 5-Dimensions 5-Levels (EQ-5D-5L) at a single time point within 30 days postpartum. Clinical data were extracted from medical records. Analyses were performed across three postpartum windows (0–2, 3–7, and 8–30 days). Structural validity, measurement invariance, reliability, and construct validity (convergent and known-groups) were assessed.

**Results:** A total of 1,935 women were included. Structural validity supported a stable four-factor structure with excellent model fit (CFI 0.995–0.997; RMSEA 0.055–0.059), and bifactor analysis supported essential unidimensionality. Measurement invariance was confirmed at metric and scalar levels across postpartum windows. Reliability was good (Cronbach’s alpha 0.83–0.86). Convergent validity was supported by moderate correlations with the EQ-5D-5L (ρ = −0.51 to −0.30), decreasing over time. Known-groups validity was demonstrated by significantly lower scores in women with poorer health status, postpartum haemorrhage, and operative or caesarean birth (all p <0.001).

**Conclusions:** The ObsQoR-10 demonstrates consistent, valid, and reliable psychometric properties for assessing postpartum recovery across the first 30 days.

## BACKGROUND

Postpartum recovery is a major public health concern, as suboptimal recovery is associated with persistent physical, psychological, and social impairments.^1,2^ Recovery refers to the process by which women regain physical health, psychological well-being, and functional capacity after childbirth, reaching a new state of balance.^3,4^ This multidimensional process evolves over time, from physical symptoms immediately after birth to broader emotional and functional challenges in the following weeks.^5,6^

This complexity makes postpartum recovery inherently difficult to assess. However, its evaluation is essential for the early identification of women at risk and to guide postnatal care ^2^. Such assessment should reflect women’s perspectives and rely on patient-reported outcome measures (PROMs).^4^ The first 30 days postpartum represent a clinically meaningful window for assessing recovery, as this period concentrates a substantial proportion of maternal readmissions and remains associated with a significant burden of physical and psychological complications.^7,8^

Although more than twenty PROMs have been used to evaluate postpartum recovery, none has been validated across the entire first postpartum month.^9^ Most instruments are validated for the early postpartum period (0–2 days)^10^ or later stages (>7 days), with few capturing the transition between these phases.^11,12^ In the absence of postpartum-specific PROMs covering this continuum, research relies predominantly on generic quality-of-life measures such as the EuroQol-5 Dimensions-5 Levels (EQ-5D-5L),^13^ which do not capture childbirth-specific domains.

The Obstetric Quality of Recovery-10 (ObsQoR-10) is widely used to assess early postpartum recovery.^14,15^ Derived from the Quality of Recovery-40 (QoR-40),^16^ it evaluates physical comfort, pain, emotional state, independence, and newborn care. The instrument has demonstrated excellent reliability and cross-cultural validity.^10,17–19^ Although its psychometric validation remains primarily restricted to the first 72 hours postpartum ^10,20^, it has also been used up to day 7 with some supporting psychometric data.^5,21^ However, comprehensive validation beyond the first three days is lacking, and its measurement properties after the first week remain unexplored.

The aim of this study was to evaluate the psychometric properties of the ObsQoR-10 across the first postpartum month.

## METHOD

We conducted a cross-sectional psychometric analysis using baseline data from the SOCRATES study,^22^ a Swiss national multilingual cohort evaluating parental health and wellbeing over the first year postpartum. Between March and June 2025, 81 maternity units (public, private, and birth centres) enrolled participants over six complete weeks. Recruitment was undertaken by trained midwives during the first postpartum week. Eligible participants were women aged ≥14 years with capacity to provide informed consent, who had given birth to one or more children (birthweight >500 g, gestational age ≥22 weeks) and who were sufficiently proficient in French, German, Italian or English. Clinical data were collected by midwives from medical records, and participants completed an online questionnaire via Research electronic data capture (REDCap®),^23^ at a self-selected postpartum time point. Questionnaires were available in French, German, Italian, and English. All participants provided electronic informed consent before data collection. Questionnaire validation adhered to COSMIN standards (COnsensus-based Standards for the selection of health Measurement INstruments)^24^ informed by a published psychometric validation framework. ^25^ Ethical approval was obtained from all seven Swiss ethics committees (ID 2024-02262), and the protocol was registered on ClinicalTrials.gov (NCT06886841).

### Study population

For this psychometric analysis, women who completed the ObsQoR-10 within 30 days postpartum were eligible. Women with stillbirth or peripartum death were excluded because two ObsQoR-10 items were not applicable (Figure 1).

**Figure 1.**
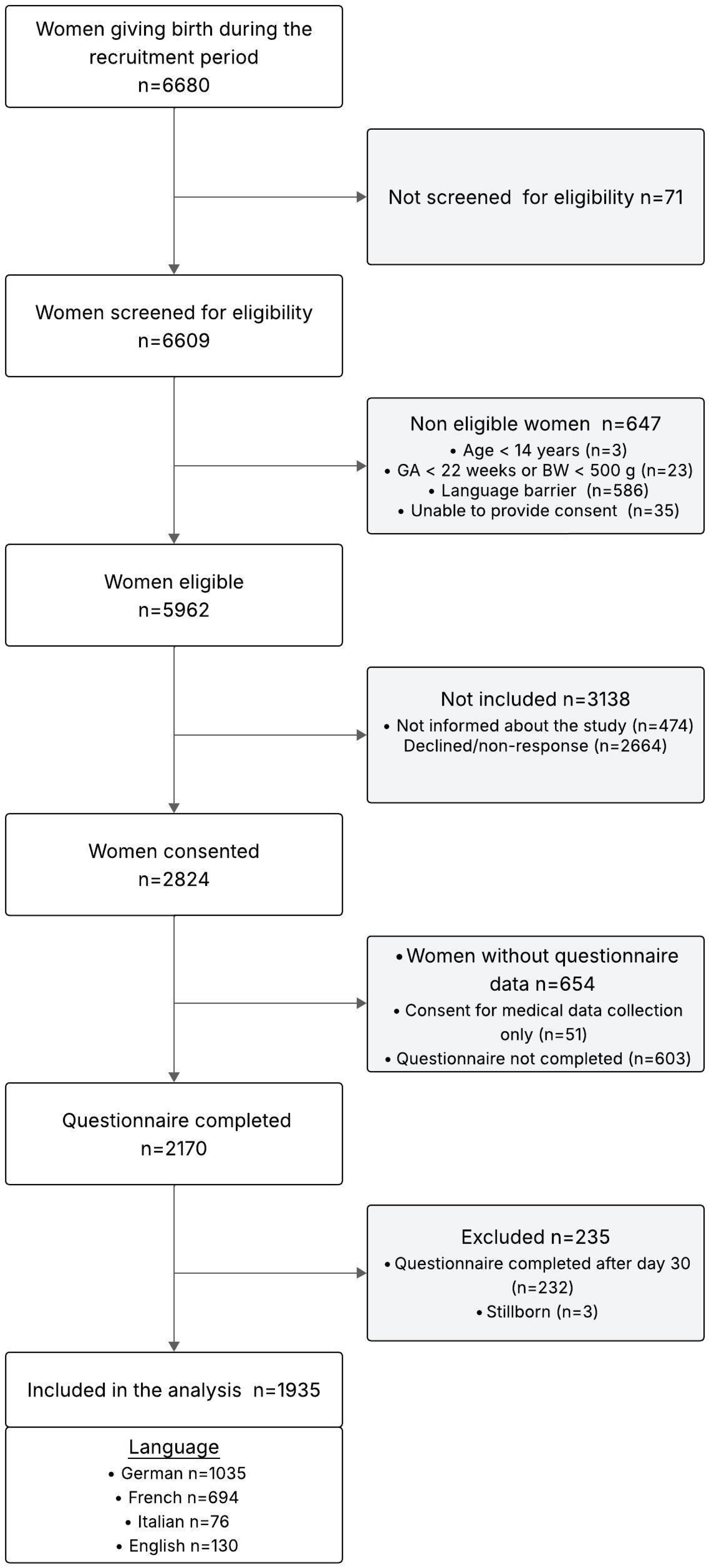
Flowchart of participant inclusion.

### Measures

Postpartum recovery was assessed using the ObsQoR-10, a 10-item PROM derived from the QoR-40.^17^ The ObsQoR-10 was adapted from the original ObsQoR-11 by merging two pain-related items.^10^ Items reflect key recovery domains, including physical comfort, autonomy, newborn care, and emotional well-being. Items are scored from 0 to 10 (total score 0–100), with higher scores indicating better recovery; negatively worded items were reverse-coded. The minimal clinically important difference is 5 points.^18^ Validated French, German, and English versions were use.^18,19^ As no Italian version existed, it was translated according to International Society for Pharmacoeconomics and Outcomes Research (ISPOR) guidelines,^26^ including forward and backward translations, expert reconciliation, and pilot testing in eight postpartum women

The EQ-5D-5L and the General Health Visual Analogue Scale (GH-VAS) were used to assess convergent validity. The EQ-5D-5L assesses health status across five domains (mobility, self-care, usual activities, pain/discomfort, and anxiety/depression), each rated on five levels.^27^ Total scores range from 5 to 25, with higher scores indicating poorer health. General health was assessed using the GH-VAS (0–100), with scores ≥70 considered indicative of good perceived health status.^28^

### Covariates

Sociodemographic and health covariates included age (<25, 25–34, ≥35 years), self-reported ethnicity (Caucasian, other), relationship status (single, couple), education level (compulsory or less, secondary, tertiary), and history of psychiatric or chronic disease.

Clinical variables included mode of childbirth (spontaneous vaginal birth, operative vaginal birth, elective or emergency caesarean delivery), parity (primiparous, multiparous), perineal trauma (intact perineum, first- or second-degree tears, third- or fourth-degree tears, episiotomy), postpartum haemorrhage (PPH ≥500 mL),^27^ Apgar score <7 at 5 minutes, ^30^ preterm birth (<37 weeks’ gestation), neuraxial labour analgesia (yes, no), neonatal intensive care transfer (yes, no) and maternal complications during postpartum stay (e.g., infection, surgical complications).

### Statistical analyses

Descriptive statistics were reported as mean and standard deviation (SD), median and quartiles (Q1–Q3) or absolute and relative frequencies (n, %). Non-parametric tests were used for between-group comparisons. Distributions of EQ-5D-5L scores and GH-VAS were described across postpartum windows to contextualise validity analyses. Missingness in covariates was minimal (<1%). Analyses were conducted on complete cases in R (version 4.4.1) ^31^ using the *psych* and *lavaan* packages.

#### Psychometric analysis

Psychometric properties were analysed across three postpartum windows: the original validation period (0–2 days), corresponding to the timeframe in which structural validity has been established ^10,18^ a subsequent period with extended use and psychometric evaluation focusing on construct validity and reliability (3–7 days) ^5,17,21^ and the largely unexplored later postpartum period within the first month (8–30 days). Analyses were also conducted within each language group using pooled data.

##### Structural validity

Data suitability for exploratory factor analysis (EFA) was assessed using Bartlett’s test and the Kaiser–Meyer–Olkin statistic. EFA was conducted separately within each postpartum window, using principal axis factoring with oblimin rotation, accounting for correlated dimensions.^32^ The number of factors was determined by parallel analysis. Because the ObsQoR-10 includes a single-item pain factor, which cannot form a standalone latent construct, it was grouped with the physical comfort items, yielding a four-factor structure consistent with previous conceptualisations.^17,20^

For comparability across postpartum windows, this four-factor solution was examined in EFA and subsequently tested using confirmatory factor analysis (CFA). CFA was conducted within each postpartum window using the weighted least squares mean and variance adjusted (WLSMV) estimator and repeated within each language group to examine structural consistency. Model fit was evaluated using comparative fit index (CFI), Tucker–Lewis index (TLI), root mean square error of approximation (RMSEA), and standardized root mean square residual (SRMR) (acceptable: CFI/TLI ≥ 0.95; RMSEA/SRMR ≤ 0.08; excellent: CFI/TLI ≥ 0.97; RMSEA ≤ 0.05).^33^

Given that the ObsQoR-10 is interpreted as a total score,^18^ a bifactor CFA model was fitted to assess whether a dominant general factor accounted for most shared variance among items. Support for treating the scale as essentially unidimensional was defined a priori according to conventional thresholds: omega hierarchical (ωH) ≥ 0.70, explained common variance (ECV) ≥ 0.60, and percentage of uncontaminated correlations (PUC) ≥ 0.80.^34^ One-factor models were also estimated for comparison. As a sensitivity analysis, split-half procedures were conducted by randomly splitting the sample into two subsamples and repeating the EFA and one- and four-factor CFA models in both subsamples.

Measurement invariance across postpartum windows was tested using sequential configural, metric, and scalar models, with invariance inferred at ΔCFI < 0.01.^35^ Items were treated as ordered categorical variables (WLSMV), and the original 11-point response scale collapsed into five ordered categories (0–2, 3–4, 5–6, 7–8, 9–10) to ensure stable estimation. Multi-group language invariance testing was not feasible due to subgroup size imbalance and parameter instability; the four-factor CFA model was therefore estimated separately within each language group.

##### Reliability

Internal consistency was assessed using Cronbach’s α, McDonald’s ω, and split-half reliability. Values ≥ 0.70, ≥ 0.80, and ≥ 0.90 were interpreted as acceptable, good, and excellent, respectively.

##### Validity

Convergent validity was assessed using Spearman correlations between ObsQoR-10 scores and EQ-5D-5L and GH-VAS. Effect sizes were interpreted as weak (|ρ| ≈ 0.10), moderate (|ρ| ≈ 0.30), or strong (|ρ| ≥ 0.50).^36^ Negative correlations with the EQ-5D-5L and positive correlations with the GH-VAS were hypothesised.

Known-groups validity was examined based on three clinically derived hypotheses: (i) women with GH-VAS < 70 would report lower ObsQoR-10 scores than those with GH-VAS ≥ 70; (ii) women with postpartum haemorrhage would have lower scores than those without; and (iii) recovery would be better after spontaneous vaginal birth than after operative vaginal or caesarean delivery. Group comparisons were performed using the Wilcoxon rank-sum test for two-group comparisons and the Kruskal–Wallis test for comparisons involving more than two groups.

##### Interpretability

Interpretability was evaluated using score distributions, inter-item correlations, and floor and ceiling effects (defined as present when >15% of respondents achieved minimum or maximum scores).^24^ Heatmaps and boxplots were used to visualise response patterns.

#### Weighting

Based on prior evidence that item salience may vary beyond the postpartum phase,^5^ we examined whether time-specific item weighting improved score precision after 2 days postpartum. Standardised general-factor loadings from bifactor CFA models were estimated across predefined postpartum windows and weekly sub-intervals within the 8–30-day period (8–14, 15–21, and 22–30 days). Temporal variation in item loadings was evaluated using mean absolute deviations to determine whether weighting should be derived from time-specific periods or from pooled postpartum windows.

Weighted scores were then compared with the original unweighted total score using Pearson correlation (≥ 0.95 indicating equivalence) and concordance at established cut-offs (≥ 77 and ≥ 86),^21^ with ≥ 90% concordance considered excellent,^37^ Weighting was considered negligible if all equivalence criteria were met.

## RESULTS

### Description of the population

Overall, 1935 women were included (Fig. 1). Of these, 206 completed the questionnaire at 0–2 days, 877 at 3–7 days, and 852 at 8–30 days postpartum. Most participants were aged 25– 34 years (62.7%), 74.5% had tertiary education, and 90.5% reported Caucasian ethnicity. Sociodemographic characteristics were similar across postpartum windows (Table 1).

**Table 1.** Participant characteristics by postpartum windows.

|  | Postpartum days |  |  |  |
| --- | --- | --- | --- | --- |
|  | 0-2<br>(N = 206) | 3-7<br>(N = 877) | 8-30<br>(N = 852) | Total<br>(N = 1935) |
| **Characteristics** |  |  |  |  |
| Age (years), n=1935 |  |  |  |  |
| < 25 | 4 (1.9) | 16 (1.8) | 12 (1.4) | 32 (1.7) |
| 25-34 | 122 (59.2) | 562 (64.1) | 530 (62.2) | 1,214 (62.7) |
| ≥35 | 80 (38.8) | 299 (34.1) | 310 (36.4) | 689 (35.6) |
| Caucasian ethnicity, n=1935 | 188 (91.3) | 788 (89.9) | 775 (91.0) | 1,751 (90.5) |
| Single mother, n=1935 | 2 (1.0) | 9 (1.0) | 13 (1.5) | 24 (1.2) |
| Education level, n=1932 |  |  |  |  |
| Compulsory or less | 5 (2.4) | 24 (2.7) | 25 (2.9) | 54 (2.8) |
| Secondary | 57 (27.7) | 211 (24.1) | 165 (19.4) | 433 (22.4) |
| Tertiary | 144 (69.9) | 641 (73.2) | 660 (77.6) | 1,445 (74.8) |
| Psychiatric history, n=1935 | 52 (25.2) | 225 (25.7) | 229 (26.9) | 506 (26.1) |
| Chronic disease, n=1932 | 90 (43.9) | 370 (42.3) | 336 (39.4) | 796 (41.2) |
| Multiparous, n=1935 | 130 (63.1) | 363 (41.4) | 288 (33.8) | 781 (40.4) |
| Multiple pregnancies, n=1935 | 0 (0) | 12 (1.4) | 10 (1.2) | 22 (1.1) |
| Preterm birth, n=1935 | 11 (5.3) | 36 (4.1) | 44 (5.2) | 91 (4.7) |
| Pregnancy complications, n=1928 | 75 (36.4) | 321 (36.8) | 309 (36.4) | 705 (36.6) |
| Mode of birth, n=1935 |  |  |  |  |
| Spontaneous vaginal birth | 142 (68.9) | 501 (57.1) | 469 (55.0) | 1,112 (57.5) |
| Operative vaginal birth | 23 (11.2) | 106 (12.1) | 113 (13.3) | 242 (12.5) |
| Elective caesarean delivery | 25 (12.1) | 121 (13.8) | 113 (13.3) | 259 (13.4) |
| Emergency caesarean delivery | 16 (7.8) | 149 (17.0) | 157 (18.4) | 322 (16.6) |
| Labor induction, n=1935 | 63 (30.6) | 263 (30.0) | 247 (29.0) | 573 (29.6) |
| Neuraxial labour analgesia, n=1933 | 122 (59.2) | 581 (66.3) | 538 (63.2) | 1,241 (64.2) |
| Perineal trauma<sup>1</sup>, n = 1353 |  |  |  |  |
| Intact perineum | 36 (21.8) | 147 (24.2) | 122 (21.0) | 305 (22.5) |
| 1<sup>st</sup> or 2<sup>nd</sup> degree tear | 112 (67.9) | 381 (62.8) | 370 (63.7) | 863 (63.8) |
| 3<sup>rd</sup> or 4<sup>th</sup> degree tear | 7 (4.2) | 20 (3.3) | 21 (3.6) | 48 (3.5) |
| Episiotomy | 10 (6.1) | 59 (9.7) | 68 (11.7) | 137 (10.1) |
| PPH, n=1935 | 58 (28.2) | 299 (34.1) | 278 (32.6) | 635 (32.8) |
| Apgar score <7 at 5 min, n=1935 | 5 (2.4) | 15 (1.7) | 27 (3.2) | 47 (2.4) |
| Neonatal intensive care transfer, n=1931 | 11 (5.3) | 48 (5.5) | 68 (8.0) | 127 (6.6) |
| Postpartum complicationS1, n=1935 | 10 (4.9) | 97 (11.1) | 106 (12.4) | 213 (11.0) |
| EQ-5D-5L, n=1928 | 8.0 [6.0- 10.0] | 8.00 [6.0- 10.0] | 7.00 [6.0- 9.0] | 8.00 [6.00- 10.0] |
| GH-VAS, n=1928 | 79 [64- 85] | 77 [66- 85] | 80 [70-88] | 79 [69-85] |
| ObsQoR-10, n=1935 | 81 [69- 89] | 83 [74-92] | 84 [69-93] | 83 [71-92] |

Categorical variables are presented as n (%), and continuous variables as median [Q1–Q3]. EQ-5D-5L: EuroQol-5 Dimensions-5 Levels; GH-VAS: General Health Visual Analogue Scale; min: minutes; ObsQoR-10: Obstetric Quality of Recovery-10; PPH: postpartum haemorrhage.
<sup>1</sup>Perineal traumas were evaluated among women with vaginal delivery only
<sup>2</sup>Postpartum complications include complications during the postpartum stay (e.g., infection, surgical complications).

Obstetric characteristics varied by questionnaire completion timing. Women responding at 0– 2 days were more frequently multiparous and had lower rates of emergency caesarean delivery and postpartum complications than those responding later postpartum. Women responding at 3–7 and 8–30 days windows had comparable obstetric profiles (Table 1).

Median ObsQoR-10 scores were similar across windows (81, 83, and 84, respectively, Table 1). EQ-5D-5L scores remained stable with right-skewed distributions and increasing floor effects (Supplementary File S1, Figures S1c–d).

## Structural validity

Factorability criteria were met across all postpartum windows (Bartlett p < 0.001; KMO 0.82–0.85). Parallel analysis suggested two factors at 0–2 days and four factors at later windows. A four-factor solution explained 59.93–64.79% of the variance (Table 2).

**Table 2.**
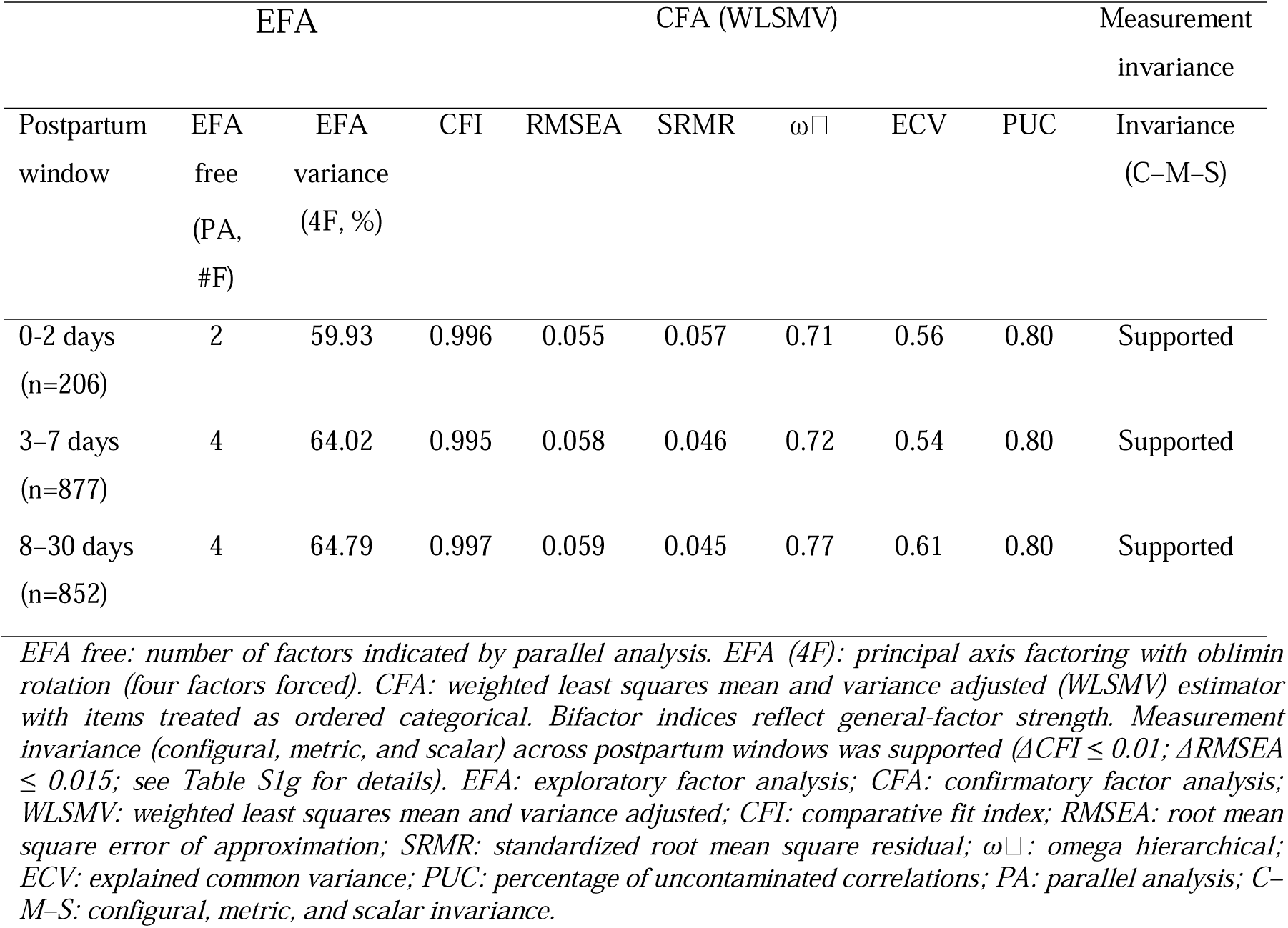
Summary of structural validity across postpartum windows.

Confirmatory analyses of the prespecified four-factor model showed excellent fit across postpartum windows (CFI 0.995–0.997; RMSEA 0.055–0.059; SRMR 0.045–0.057), with standardized loadings from 0.57 to 1.00. Bifactor indices were ωH 0.71–0.77, ECV 0.54–0.61, and PUC 0.80, consistent with essential unidimensionality.

One-factor models did not meet the predefined fit criteria; however, exploratory one-factor EFA showed acceptable item loadings (> 0.32). Multi-group CFA met thresholds for configural, metric, and scalar invariance across postpartum windows (ΔCFI ≤ 0.010; ΔRMSEA ≤ 0.015). Comparable fit was observed across language groups, and split-half analyses confirmed structural robustness (Supplementary File S2, Tables S2a–g and Figure S2).

### Reliability

Internal consistency was good across postpartum windows (α 0.83–0.86; ω 0.81–0.85; split-half 0.84–0.87; Table 3) and comparable across languages (α 0.84–0.88; ω 0.83–0.88; split-half 0.86–0.88; Supplementary File S3, Table S3a).

**Table 3.** Reliability indices of the ObsQoR-10 across postpartum windows.

| Postpartum window | Internal consistency |  | Split-half reliability |
| --- | --- | --- | --- |
| | Cronbach's $\alpha$ | McDonald's $\omega$ (95% CI) | Spearman–Brown coefficient |
| 0-2 days (n=206) | 0.83 | 0.81 (0.74–0.86) | 0.84 |
| 3–7 days (n=877) | 0.85 | 0.85 (0.81–0.86) | 0.86 |
| 8–30 days (n=852) | 0.86 | 0.85 (0.83–0.87) | 0.87 |

### Construct Validity

ObsQoR-10 scores were strongly correlated with EQ-5D-5L at 0–2 days and moderately thereafter, while correlations with GH-VAS were moderate across all postpartum windows (all Spearman p < 0.001; Table 4). Correlation magnitudes were comparable across languages.

**Table 4.** Convergent and known-groups validity of the ObsQoR-10 across postpartum windows.

| Postpartum window | Convergent validity |  | GH-VAS |  | PPH |  | Mode of birth |  |
| --- | --- | --- | --- | --- | --- | --- | --- | --- |
| | $\rho$ (EQ-5D-5L) | $\rho$ (GH-VAS) | Good health status $\geq 70$ | Poor health status $< 70$ | No PPH | PPH | SVB | Operative vaginal birth/CD |
| 0-2 days | -0.51 | 0.39 | 83 [75-92] | 70 [60-83] | 83 [72-92] | 71 [59-83] | 84 [76-92] | 70 [57-81] |
| 3-7 days | -0.48 | 0.39 | 87 [78-93] | 76 [61-85] | 86 [77-92] | 79 [64-88] | 88 [78-93] | 79 [64-88] |
| 8-30 days | -0.30 | 0.28 | 86 [71-94] | 76 [60-86] | 85 [72-93] | 79 [59-92] | 86 [76-94] | 77 [60-90] |

Values are medians [Q1–Q3] of ObsQoR-10 total scores. Analyses were conducted using complete cases; sample sizes varied slightly due to missing data. Spearman correlations and non-parametric tests (Wilcoxon rank-sum and Kruskal–Wallis) were used. All correlations and group comparisons were statistically significant ( $p < 0.001$ ).
EQ-5D-5L: EuroQol-5 Dimensions-5 Levels; GH-VAS: General Health Visual Analogue Scale; PPH: postpartum haemorrhage; SVB: spontaneous vaginal birth; CD, caesarean delivery.

For known-groups validity, ObsQoR-10 scores were consistently lower among women with poorer health status (GH-VAS <70), postpartum haemorrhage, and operative or caesarean delivery compared with their respective reference groups, across all postpartum windows (all p ≤ .001; Table 4). Similar patterns were observed across language groups (Supplementary File S3, Tables S3b–c).

### Interpretability

ObsQoR-10 scores were right-skewed, with low but increasing ceiling proportions across postpartum windows (0.5–3.3%). Item-level medians and interquartile ranges were stable across windows. Significant between-window differences were observed for several items, notably pain, nausea, dizziness, and shivering (Figure 2). Additional interpretability indices (item-level ceiling proportions, inter-item correlation heatmaps, and full score distributions) are in Supplementary File S1 (Tables S1a–b; Figures S1a–f).

**Figure 2.**
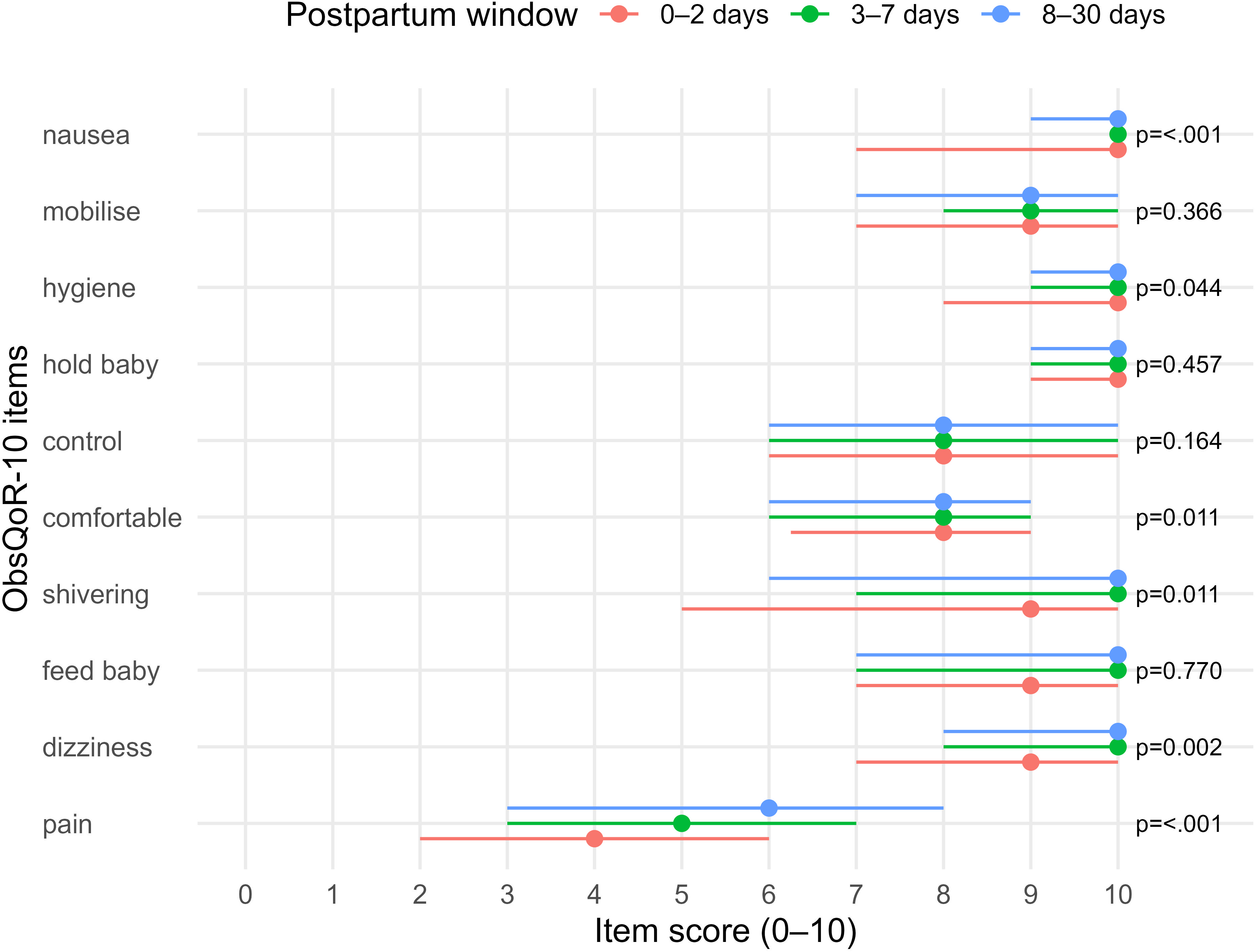
Item-level medians and interquartile ranges of ObsQoR-10 scores across postpartum windows.

### Weighting

General-factor loadings showed larger deviations from the pooled 3–30-day estimates at 0-2 days (mean |Δ| = 0.075), whereas deviations were minimal across 3–7 days, 8–30 days, and across weekly sub-intervals within the 8–30-day period (8–14 days, n=516; 15–21 days, n=215; 22–30 days, n=121) (mean |Δ| = 0.014–0.034). Item weights were therefore derived from the pooled 3–30-day model.

In the 8–30-day window, weighted and unweighted ObsQoR-10 scores showed near-perfect agreement (Pearson r = 0.979; Spearman ρ = 0.985) and reclassification at the ≥77 and ≥86 cut-offs was low (5.9% and 4.7%, respectively), indicating negligible impact on score interpretation. Additional weighting diagnostics are provided in Supplementary File S4 (Tables S4a–d).

## DISCUSSION

In this study, the ObsQoR-10 demonstrated consistent psychometric performance across the first 30 days postpartum and discriminated between clinically relevant groups. Correlations with the EQ-5D-5L were strong in the early postpartum period and decreased to moderate levels over time.

Consistent with early postpartum validations,^10,17^ the ObsQoR-10 retained a multidimensional four-factor structure across postpartum windows. Although exploratory analyses suggested a transient two-factor pattern within the first two days postpartum, confirmatory analyses supported the predefined model in all windows. This early pattern likely reflects clustering of acute symptoms rather than a distinct latent construct. Bifactor analyses supported essential unidimensionality, justifying use of the global score and extending earlier findings of a dominant first factor.^18^ Minor temporal changes in item behavior, including small variations in loadings and increasing ceiling effects for shivering, dizziness, and holding the baby, were observed but did not affect interpretability. These changes are consistent with expected recovery trajectories, as acute symptoms and functional limitations typically resolve within the first days postpartum.^5,6^

In addition, the ObsQoR-10 demonstrated sustained discriminative validity across windows, distinguishing between mode of birth, postpartum haemorrhage and general health status, with differences exceeding the minimal clinically important difference.^18^ This indicates that the score retained sensitivity to differences in recovery beyond the first two days. The ObsQoR-10 captured heterogeneity in recovery during the later postpartum period, with first-quartile scores between 8 and 30 days postpartum remaining below the clinically referenced threshold of 77.^21^ Similar score distributions have been reported previously, with wide dispersion at discharge and during the first postpartum week.^21,38^ Together, these findings support the ObsQoR-10’s ability to discriminate recovery profiles beyond the early postpartum period.

Finally, the evolving correlation between the ObsQoR-10 and the EQ-5D-5L may support the specificity of the recovery construct captured by the ObsQoR-10. Correlations were moderate in the early postpartum and attenuated over time, consistent with prior reports ^13^. This pattern aligns with the conceptual distinction between postpartum recovery, which remains dynamic over the first month, and generic health-related quality of life, which tends to stabilise once major morbidity has resolved.^39^ While EQ-5D-5L scores approached optimal values after the first week, the ObsQoR-10 retained variability consistent with known clinical recovery patterns,^7^ suggesting greater sensitivity to postpartum recovery trajectories.

Clinically, these findings indicate that the ObsQoR-10 can provide a stable overview of recovery across the first postpartum month. Although the ObsQoR-10 captures key dimensions of early postpartum recovery, it was not designed to provide a comprehensive assessment beyond the three days after childbirth.^17^ From day three postpartum, domains such as fatigue, sleep quality, and psychosocial adaptation may become increasingly salient and are only partially represented in the scale.^5,40^ In this context, the ObsQoR-10 reflects overall recovery rather than a comprehensive assessment of postpartum health beyond the first days after childbirth. When used for routine assessment during the first 30 days postpartum, the scale may therefore be complemented by broader instruments, such as the Stanford Obstetric Recovery Checklist (STORK).^11^

This study has several strengths. A comprehensive psychometric framework enabled the evaluation of structural validity and temporal stability across postpartum windows. The large, nationally recruited cohort allowed assessment across multiple language versions within a unified analytical framework. Comparable model fit, reliability, and construct validity across translations suggest stable psychometric performances, although formal multi-group invariance testing was not conducted. Comparison with the EQ-5D-5L contextualized convergent validity and highlighted the specificity of obstetric recovery constructs.

Several limitations should also be acknowledged. Each participant completed the ObsQoR-10 at a single postpartum time point, precluding assessment of individual recovery trajectories and responsiveness to change. Accordingly, comparisons across postpartum windows reflect psychometric performance at different time points rather than longitudinal recovery. Weighting analyses suggested stable psychometric performance across weekly postpartum subgroups; however, smaller subgroup sizes in the later windows may have reduced estimate precision. Finally, while language-specific confirmatory analyses were conducted, formal multi-group language invariance testing was not undertaken. Independent evaluation of the Italian version in a larger dedicated sample is warranted.

Future research should further characterize postpartum recovery trajectories and refine clinical application of the ObsQoR-10 across the first postpartum month. Longitudinal studies are needed to assess within-individual trajectories, responsiveness, test–retest reliability, and predictive validity of early postpartum scores to inform recovery monitoring and postpartum care.

## CONCLUSION

This study demonstrates that the ObsQoR-10 exhibits consistent and valid psychometric properties across the first 30 days postpartum. Future longitudinal studies should examine recovery trajectories and further clarify the usefulness of the ObsQoR-10 within postpartum care pathways.

## Data Availability

All data produced in the present study are available upon reasonable request to the authors

## CRediT authorship contribution statement

LG and TD contributed equally as last authors.

EC: Conceptualisation, Investigation, Methodology, Data curation, Validation, Formal analysis, Writing – original draft, Writing – review & editing.

EL: Conceptualisation, Investigation, Methodology, Project administration, Writing – review & editing.

GC: Data curation, Methodology, Validation, Writing – review & editing.

ME: Investigation, Writing – review & editing.

SGB: Funding acquisition, Project administration, Writing – review & editing.

ANM: Investigation, Data curation, Writing – review & editing.

JS: Writing – review & editing.

MS: Writing – review & editing.

INW: Investigation, Data curation, Writing – review & editing.

LG: Conceptualisation, Methodology, Funding acquisition, Project administration, Supervision, Writing – review & editing.

TD: Conceptualisation, Methodology, Supervision, Writing – review & editing.

All authors critically reviewed and approved the final version of the manuscript.

## Acknowledgements

The authors would like to thank all participating maternity units, birth centres, and midwifery organisations across Switzerland for their essential contribution to data collection. We are grateful to the partner institutions and organisations involved in the SOCRATES study for their support in the development and implementation of the project. We also acknowledge all members of the SOCRATES study team, as well as the healthcare professionals involved in participant recruitment and local data management. We sincerely thank the parents who generously contributed their time to this study.

We further thank Alexandra L. Dima for her valuable methodological insights and guidance.

## Declaration of generative AI and AI-assisted technologies in the manuscript preparation process

During the preparation of this work, the authors used ChatGPT (OpenAI) and NotebookLM (Google) in order to assist with language editing and improve the clarity of the manuscript. After using these tools, the authors reviewed and edited the content as needed and take full responsibility for the content of the published article.

