## Supplementary File S4 for "Psychometric validation of the Obstetric Quality of Recovery-10 scoring tool across the first month postpartum: a cross-sectional psychometric study"

### **Supplementary material S5: Weighting**

General-factor loadings from bifactor confirmatory factor analysis (CFA) models were compared across postpartum windows to assess the stability of item contributions and the appropriateness of weighted scoring. Loadings showed greater variability in the 0–2 days and 22–30 days windows, whereas differences were minimal across the 3–7 days and 8–30 days periods. Item weights were therefore derived from the pooled 3–30 days model and normalized to sum to 10. Weighted and unweighted total scores were compared using Pearson and Spearman correlations and agreement at established ObsQoR-10 cut-offs.

| Item | 0-2 days  n=206 | 3–7 days  n=877 | 8–30 days  n=852 | 3–30 days  n=1729 | 8–14 days  n=516 | 15–21 days  n=215 | 22–30 days  n=121 |
| --- | --- | --- | --- | --- | --- | --- | --- |
| Comfortable | 0.58 | 0.59 | 0.61 | 0.60 | 0.61 | 0.60 | 0.62 |
| Control | 0.91 | 0.83 | 0.84 | 0.83 | 0.85 | 0.77 | 0.87 |
| Dizziness | 0.51 | 0.37 | 0.41 | 0.39 | 0.38 | 0.43 | 0.52 |
| Feed baby | 0.53 | 0.70 | 0.72 | 0.71 | 0.75 | 0.66 | 0.70 |
| Hold baby | 0.77 | 0.84 | 0.79 | 0.81 | 0.76 | 0.79 | 0.91 |
| Hygiene | 0.88 | 0.89 | 0.94 | 0.92 | 0.95 | 0.95 | 0.93 |
| Mobilisation | 0.89 | 0.86 | 0.91 | 0.89 | 0.91 | 0.93 | 0.92 |
| Nausea | 0.32 | 0.34 | 0.34 | 0.34 | 0.35 | 0.29 | 0.45 |
| Pain | 0.27 | 0.44 | 0.49 | 0.47 | 0.48 | 0.51 | 0.53 |
| Shivering | 0.33 | 0.34 | 0.41 | 0.38 | 0.39 | 0.39 | 0.53 |
| *Values represent standardized general-factor loadings (est.std) from bifactor CFA models estimated using the WLSMV estimator. Items were treated as ordered categorical variables. CFA: confirmatory factor analysis; WLSMV: weighted least squares mean and variance adjusted.* | | | | | | | |

**Table S5a. Bifactor general-factor loadings (G) across postpartum windows**

#### **Table S5b. Mean absolute deviation (|Δ|) of general-factor loadings relative to the pooled 3–30 days model.**

| Postpartum windows | Mean \|Δ\| |
| --- | --- |
| 0-2 days (n=206) | 0.075 |
| 3–7 days (n=877) | 0.019 |
| 8–30 days (n=852) | 0.016 |
| 8–14 days (n=516) | 0.020 |
| 15–21 days (n=215) | 0.035 |
| 22–30 days (n=121) | 0.066 |

*Values represent the mean absolute difference between window-specific standardised general-factor loadings and those estimated from the pooled 3–30 days bifactor model. Lower values indicate greater stability of item loadings across time.*

**Table S5c. Bifactor-based general-factor loadings and item weights (3–30 days)**

| Items | G loading (standardised) | Normalised weight (sum = 10) |
| --- | --- | --- |
| Comfortable | 0.60 | 0.94 |
| Control | 0.83 | 1.31 |
| Dizziness | 0.39 | 0.61 |
| Feed baby | 0.71 | 1.12 |
| Hold baby | 0.81 | 1.28 |
| Hygiene | 0.92 | 1.45 |
| Mobilisation | 0.89 | 1.40 |
| Nausea | 0.34 | 0.54 |
| Pain | 0.47 | 0.74 |
| Shivering | 0.38 | 0.60 |

#### **Table S5d. Agreement of weighted vs unweighted scores (3–30 days)**

| Metric | Value |
| --- | --- |
| Pearson correlation (r) | 0.979 |
| Spearman correlation (ρ) | 0.985 |
| Median score difference (weighted − unweighted) | 0.55 |
| Reclassification at cut-off ≥ 77 | 5.8% |
| Reclassification at cut-off ≥ 86 | 4.6% |
| *Weighted scores were derived from bifactor general-factor loadings estimated on the pooled 3–30-day sample. Agreement was assessed using correlation coefficients, absolute score differences, and concordance at established ObsQoR-10 cut-offs (≥ 77 and ≥ 86).* | |
