## Supplementary File S3 for "Psychometric validation of the Obstetric Quality of Recovery-10 scoring tool across the first month postpartum: a cross-sectional psychometric study"

### **Supplementary material S4: Construct validity**

#### **Table S4a. Reliability indices of the ObsQoR-10 by language**

| Language | Cronbach’s α | McDonald’s ω | Split-half (Spearman–Brown) |
| --- | --- | --- | --- |
| French (n=694) | 0.86 | 0.85 (95% CI 0.83–0.88) | 0.87 |
| German (n=1035) | 0.85 | 0.83 (95% CI 0.80–0.85) | 0.86 |
| Italian (n=76) | 0.84 | 0.86 (95% CI 0.77–0.91) | 0.86 |
| English (n=130) | 0.88 | 0.87 (95% CI 0.80–0.91) | 0.88 |
| *Values are point estimates; McDonald’s ω includes 95% confidence intervals. Split-half reliability was estimated using the Spearman–Brown coefficient. Bifactor indices were not computed at the language level due to limited sample size, particularly for the Italian subgroup.* | | | |

#### **Table S4b. Known-group validity of the ObsQoR-10 by language**

|  | | GH-VAS | | PPH | | | | Mode of birth | | | | |
| --- | --- | --- | --- | --- | --- | --- | --- | --- | --- | --- | --- | --- |
| Language | ≥70 | | <70 | | p | No PPH | PPH | | p | SVB | Operative vaginal birth/CD | p |
| French (n=694) | 88 [78-93] | | 77 [64-86] | | <0.001 | 87 [77-92] | 80 [62-88] | | <0.001 | 88 [78-93] | 80 [63-89] | <0.001 |
| German (n=1035) | 84 [73-93] | | 75 [58-86] | | <0.001 | 84 [74-93] | 79 [63-90] | | <.0001 | 86 [77-94] | 76 [60-88] | <0.001 |
| Italian (n=76) | 83 [73-92] | | 72 [57-82] | | 0.012 | 82 [77-92] | 75 [57-84] | | 0.036 | 82 [76-88] | 79 [57-92] | 0.130 |
| English (n=130) | 87 [74-95] | | 72 [59-86] | | <0.001 | 85[70-94] | 82 [61-90] | | 0.200 | 85 [73-96] | 79 [61-90] | 0.110 |

#### *Values are medians [Q1-Q3] of ObsQoR-10 total scores. Group comparisons were performed using Wilcoxon rank-sum tests. GH-VAS, General Health Visual Analogue Scale; PPH, postpartum haemorrhage; SVB, spontaneous vaginal birth; CD, caesarean delivery.*

#### **Table S4c. Convergent validity of the ObsQoR-10 by language**

|  | Convergent validity | | | |
| --- | --- | --- | --- | --- |
| Language | ρ (EQ-5D-5L) | p | ρ (GH-VAS) | p |
| French (n=694) | -0.37 | <0.001 | 0.36 | <0.001 |
| German (n=1035) | -0.37 | <0.001 | 0.29 | <0.001 |
| Italian (n=76) | -0.49 | <0.001 | 0.38 | 0.001 |
| English (n=130) | -0.45 | <0.001 | 0.46 | <0.001 |

*Spearman correlations were used to assess convergent validity. EQ-5D-5L, EuroQol-5 Dimensions-5 Levels; GH-VAS, General Health Visual Analogue Scale.*
