## Supplementary File S2 for "Psychometric validation of the Obstetric Quality of Recovery-10 scoring tool across the first month postpartum: a cross-sectional psychometric study"

**Supplementary material S3: Structural validity**

**Table S3b. CFA Fit indices by postpartum window and language**

| Postpartum window | |  |  | |  | | |  | Model fit indices | | | | | | | |
| --- | --- | --- | --- | --- | --- | --- | --- | --- | --- | --- | --- | --- | --- | --- | --- | --- |
|  |  | χ² | df | | χ²/df | | | p | CFI | | TLI | | RMSEA | | SRMR | |
| 0-2 days (n=206) | 46.98 | | | 29 | | 1.62 | 0.019 | | | 0.996 | | 0.994 | | 0.055 | | 0.057 |
| 3–7 days (n=877) | 114.29 | | | 29 | | 3.94 | <0.001 | | | 0.995 | | 0.993 | | 0.058 | | 0.046 |
| 8–30 days (n=852) | 114.19 | | | 29 | | 3.94 | <0.001 | | | 0.996 | | 0.996 | | 0.059 | | 0.045 |
| Language | | | | | | | | | | | | | | | | |
| French (n=694) | 150.04 | | | 29 | | 5.17 | <0.001 | | | 0.992 | | 0.988 | | 0.078 | | 0.055 |
| German (n=1035) | 132.10 | | | 29 | | 4.56 | <0.001 | | | 0.996 | | 0.994 | | 0.059 | | 0.050 |
| Italian (n=76) | 27.15 | | | 29 | | 0.94 | 0.564 | | | 1.000 | | 1.001 | | 0.000 | | 0.072 |
| English (n=130) | 49.68 | | | 29 | | 1.71 | 0.010 | | | 0.996 | | 0.993 | | 0.074 | | 0.055 |
| *WLSMV. Items were treated as ordered categorical variables. Model fit was evaluated using χ², χ²/df, CFI, TLI, RMSEA, and SRMR. χ²: chi-square statistic; df: degrees of freedom; CFI: comparative fit index; TLI: Tucker–Lewis index; RMSEA: root mean square error of approximation; SRMR: standardized root mean square residual; WLSMV: weighted least squares mean and variance adjusted.* | | | | | | | | | | | | | | | | |

**Table S3c: Standardized CFA factor loadings by item across postpartum windows and language versions**

| Domain | Item | 0-2 days  (n=206) | 3–7 days  (n=877) | 8–30 days  (n=852) | French  (n=694) | German  (n=1035) | Italian  (n=76) | English  (n=130) |
| --- | --- | --- | --- | --- | --- | --- | --- | --- |
| Physical comfort | Dizziness | 0.79 | 0.74 | 0.69 | 0.80 | 0.65 | 0.72 | 0.81 |
|  | Nausea | 0.73 | 0.83 | 0.77 | 0.83 | 0.76 | 0.81 | 0.86 |
|  | Pain | 0.57 | 0.73 | 0.77 | 0.65 | 0.81 | 0.77 | 0.83 |
|  | Shivering | 0.72 | 0.75 | 0.79 | 0.76 | 0.77 | 0.69 | 0.81 |
| Emotional state | Comfortable | 0.59 | 0.63 | 0.63 | 0.69 | 0.60 | 0.59 | 0.61 |
|  | Control | 0.93 | 0.90 | 0.87 | 0.89 | 0.90 | 0.94 | 0.88 |
| Physical independence | Hygiene | 0.90 | 0.90 | 0.96 | 0.90 | 0.94 | 0.98 | 0.89 |
|  | Mobilisation | 0.91 | 0.87 | 0.92 | 0.87 | 0.93 | 0.89 | 0.83 |
| Care of newborn | Feed baby | 0.64 | 0.80 | 0.83 | 0.80 | 0.80 | 0.72 | 0.84 |
|  | Hold baby | 0.94 | 0.96 | 0.91 | 0.95 | 0.92 | 1.03 | 0.93 |

*Standardized CFA factor loadings from a four-domain ObsQoR-10 model (WLSMV estimator; ordered categorical items). Values indicate factor loadings and may exceed 1.0 due to the estimation method and small subgroup sample sizes. CFA: confirmatory factor analysis; WLSMV: weighted least squares mean and variance adjusted.*

**Table S3d. One-factor (unidimensional) model results across postpartum windows**

| Postpartum window | EFA variance explained (%) | CFI (1-factor model) | RMSEA (1-factor model) | SRMR (1-factor model) |
| --- | --- | --- | --- | --- |
| 0-2 days (n=206) | 35.5 | 0.957 | 0.170 | 0.127 |
| 3–7 days (n=877) | 38.6 | 0.942 | 0.184 | 0.146 |
| 8–30 days (n=852) | 40.3 | 0.965 | 0.172 | 0.133 |

### *One-factor (unidimensional) model results from EFA and CFA. Values represent variance explained and model fit indices. EFA: exploratory factor analysis; CFA: confirmatory factor analysis; CFI: comparative fit index; RMSEA: root mean square error of approximation; SRMR: standardized root mean square residual.*

**Table S3e: Split-sample one-factor EFA and CFA results across postpartum windows**

| Postpartum window | EFA one-factor variance (%) | CFI (1-factor CFA) | RMSEA (1-factor CFA) | SRMR (1-factor CFA) |
| --- | --- | --- | --- | --- |
| 0-2 days (n=206) | 41.1 | 0.926 | 0.192 | 0.152 |
| 3–7 days (n=877) | 38.2 | 0.940 | 0.184 | 0.143 |
| 8–30 days (n=852) | 42.7 | 0.957 | 0.170 | 0.133 |
| *One-factor models were estimated on split samples to assess the adequacy of a unidimensional solution. Values represent variance explained and model fit indices.*  *EFA: exploratory factor analysis; CFA: confirmatory factor analysis; CFI: comparative fit index; RMSEA: root mean square error of approximation; SRMR: standardized root mean square residual.* | | | | |

**Table S3f: Split-half evaluation of the four-factor ObsQoR-10 structure**

| Postpartum window | EFA 4F variance (%) – full sample | EFA 4F variance (%) – split-half | CFA CFI (split-half) | CFA RMSEA (split-half) | CFA SRMR (split-half) |
| --- | --- | --- | --- | --- | --- |
| 0-2 days (n=206) | 64.07 | 68.11 | 0.985 | 0.094 | 0.093 |
| 3–7 days (n=877) | 63.79 | 62.03 | 0.993 | 0.070 | 0.056 |
| 8–30 days (n=852) | 64.70 | 67.07 | 0.996 | 0.055 | 0.049 |

*Four-factor models were estimated using split-half samples to assess structural robustness. Values representvariance explained (EFA) and model fit indices (CFA).*

*EFA: exploratory factor analysis; CFA: confirmatory factor analysis; CFI: comparative fit index; RMSEA: root mean square error of approximation; SRMR: standardized root mean square residual; WLSMV: weighted least squares mean and variance adjusted.*

| Model | χ² | df | p | CFI | TLI | RMSEA | SRMR | ΔCFI | ΔRMSEA | Criteria met |
| --- | --- | --- | --- | --- | --- | --- | --- | --- | --- | --- |
| Configural | 236.65 | 87 | <0.001 | 0.996 | 0.994 | 0.052 | 0.051 | — | — | — |
| Metric | 271.90 | 99 | <0.001 | 0.996 | 0.994 | 0.052 | 0.054 | -0.001 | 0.000 | Yes |
| Scalar | 303.23 | 151 | <0.001 | 0.996 | 0.997 | 0.040 | 0.052 | 0.000 | -0.013 | Yes |
| *Measurement invariance was assessed using sequential configural, metric, and scalar models. Invariance was supported when \|ΔCFI\| ≤ 0.010 and \|ΔRMSEA\| ≤ 0.015.*  *Items were treated as ordered categorical variables using the WLSMV estimator; the original 11-point scale was collapsed into five ordered categories for invariance testing. χ²: chi-square statistic; df: degrees of freedom; CFI: comparative fit index; TLI: Tucker–Lewis index; RMSEA: root mean square error of approximation; SRMR: standardized root mean square residual; WLSMV: weighted least squares mean and variance adjusted.* | | | | | | | | | | |

**Table S3g. Measurement invariance across postpartum windows (configural, metric, scalar models)**
