## Supplementary File S1 for "Psychometric validation of the Obstetric Quality of Recovery-10 scoring tool across the first month postpartum: a cross-sectional psychometric study"

### **Supplementary material S2: Interpretability**

#### **Table S2a. Total-score distribution by postpartum window and language**

|  | By postpartum window | | | | | By language | | | | |
| --- | --- | --- | --- | --- | --- | --- | --- | --- | --- | --- |
| Characteristic | 0-2 days (n=206) | 3-7  days (n=877) | 8-30  days (n=852) | Total (n = 1935) | p-value | English (n=130) | French (n=694) | German (n=1035) | Italian (n=76) | p-value |
| Mean ± SD | 77.33 ± 15.62 | 79.89 ± 15.98 | 78.83 ± 17.56 | 79.07± 16.77 | 0.038 | 78.45 ± 17.42 | 80.42 ± 16.14 | 78.46 ± 16.98 | 76.14 ± 17.89 | 0.046 |
| Median | 81 | 83 | 84 | 83 |  | 83 | 85 | 82 | 81 |  |
| Min | 21 | 9 | 17 | 9 |  | 13 | 9 | 15 | 28 |  |
| Max | 100 | 100 | 100 | 100 |  | 100 | 100 | 100 | 100 |  |
| Q1 | 69 | 74 | 69 | 71 |  | 68 | 74 | 70 | 64 |  |
| Q3 | 89 | 92 | 93 | 92 |  | 92 | 92 | 92 | 90 |  |
| Floor n (%) | 0 (0.0%) | 0 (0.0%) | 0 (0.0%) | 0 (0.0%) |  | 0 (0.0%) | 0 (0.0%) | 0  (0.0%) | 0 (0.0%) |  |
| Ceiling n (%) | 1 (0.5%) | 9 (1.0%) | 28 (3.3%) | 38 (2.0%) |  | 6 (5.8%) | 9 (1.3%) | 21 (2.0%) | 1 (1.3%) |  |

#### *p-values were derived from Kruskal–Wallis tests comparing ObsQoR-10 total scores across collection periods (0–2, 3–7, and 8–30 days) and language groups.*

#### **Table S2b. Item-level ceiling proportions by postpartum window and language**

|  | Ceiling effect by window (%) | | | Ceiling effect by language (%) | | | |
| --- | --- | --- | --- | --- | --- | --- | --- |
| Item | 0-2 days (n=206) | 3–7 days (n=877) | 8–30 days (n=852) | English (n=130) | French (n=694) | German (n=1035) | Italian (n=76) |
| Comfortable | 22.3 | 14.8 | 20.1 | 16.2 | 14.7 | 20.5 | 15.8 |
| Control | 35.9 | 34.4 | 32.6 | 32.3 | 38.0 | 31.3 | 31.6 |
| Dizziness | **45.1** | **58.0** | **58.9** | **54.6** | **63.7** | **52.5** | **63.2** |
| Feed baby | **49.5** | **52.3** | **52.9** | **44.6** | **54.8** | **52.2** | **44.7** |
| Hold baby | **67.0** | **71.2** | **68.8** | **56.2** | **69.0** | **72.7** | **57.9** |
| Hygiene | **64.6** | **71.5** | **68.3** | **60.8** | **73.6** | **69.0** | **50.0** |
| Mobilisation | **44.2** | **47.9** | **49.1** | **35.4** | **48.1** | **50.0** | **42.1** |
| Nausea | **62.1** | **75.5** | **73.7** | **72.3** | **79.5** | **69.8** | **65.8** |
| Pain | 1.5 | 3.6 | 9.2 | 9.2 | 5.2 | 5.9 | 5.3 |
| Shivering | **45.6** | **57.8** | **57.2** | **60.0** | **62.1** | **51.6** | **59.2** |
| *Ceiling = % with item score = 10. Bold indicates ≥50% in any column.* | | | | | | | |

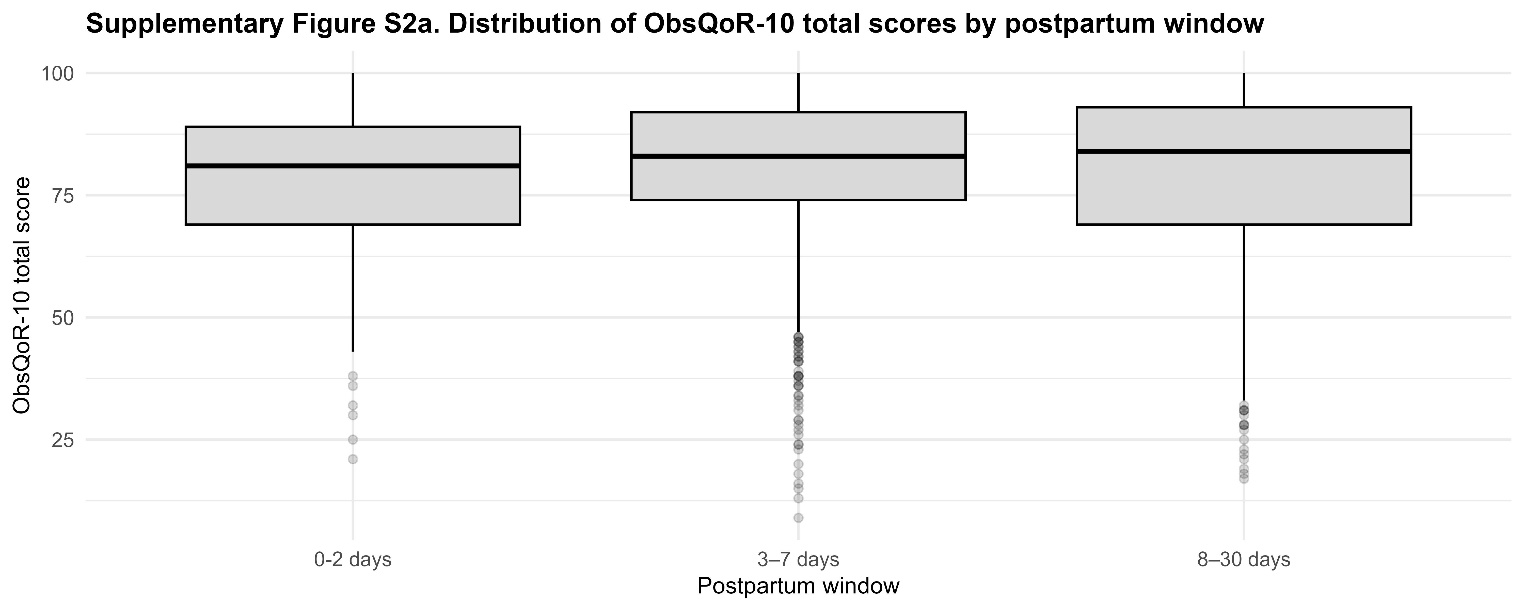

#### **Figure S2a. ObsQoR-10 distributions by postpartum window**

*Boxplots display the median, interquartile range (IQR), and range. Points represent individual observations.*

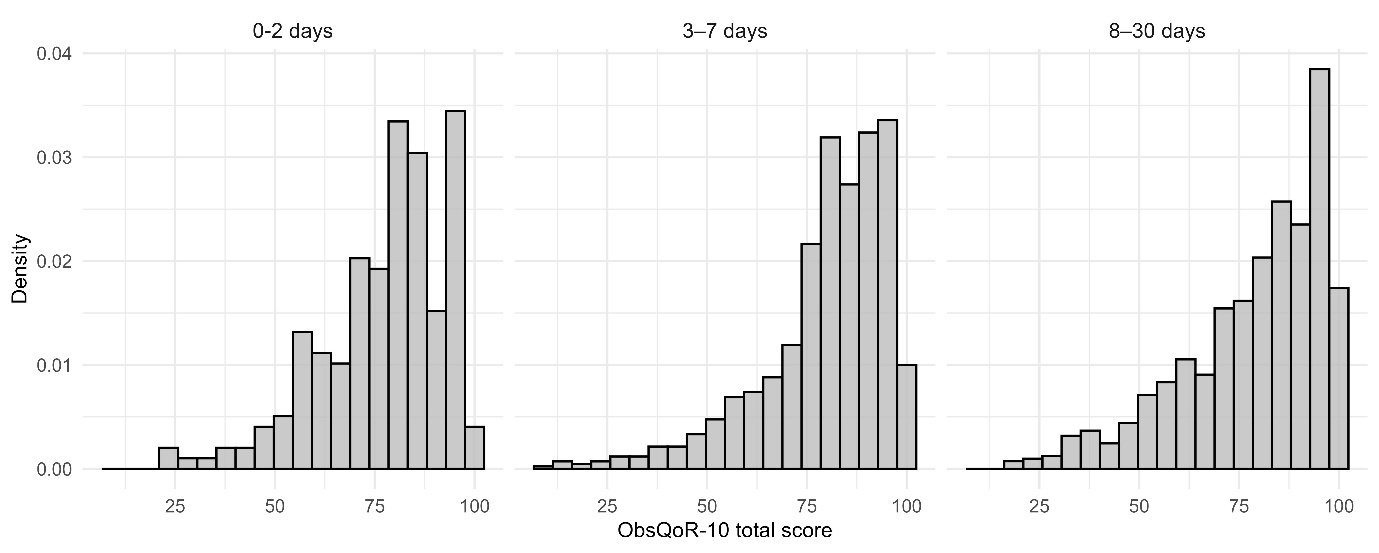

**Figure S2b. Histogram of ObsQoR-10 total scores by postpartum window**

*Histograms display the distribution of ObsQoR-10 scores across postpartum windows. Density is shown on the y-axis. Higher scores indicate better recovery.*

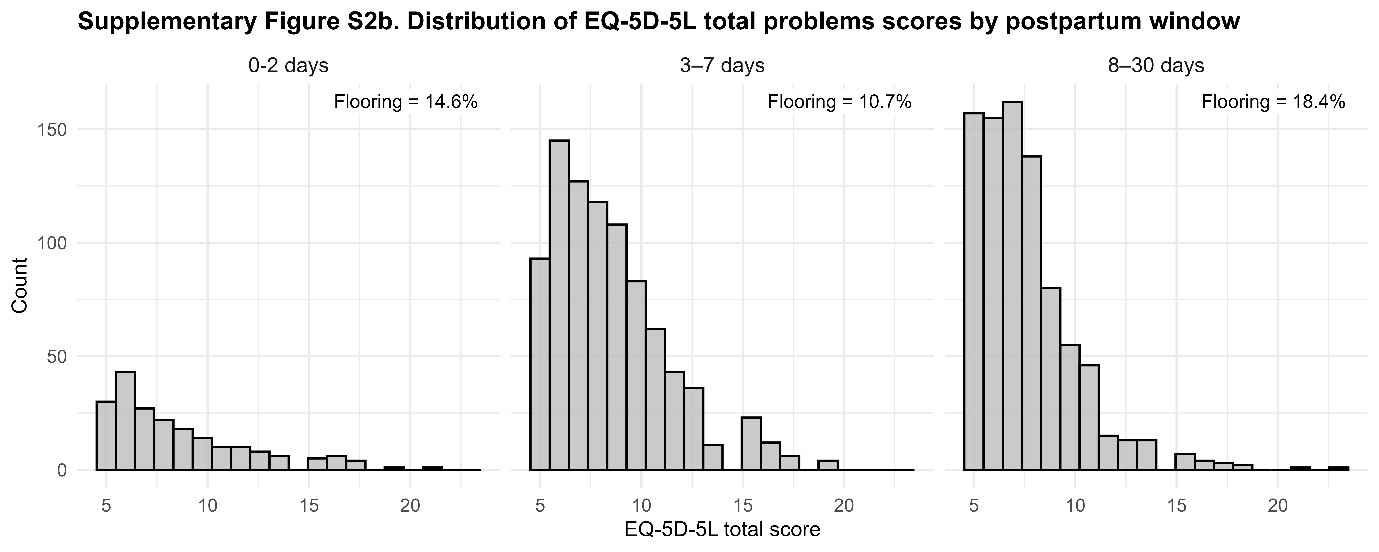

#### **Figure S2c. Histograms of EQ-5D-5L scores by postpartum window, with corresponding flooring percentages**

*Histograms display the distribution of EQ-5D-5L total scores across postpartum windows. Counts are shown on the y-axis. Flooring effects are indicated for each time window, with increasing proportions over time. Higher scores indicate poorer health status.*

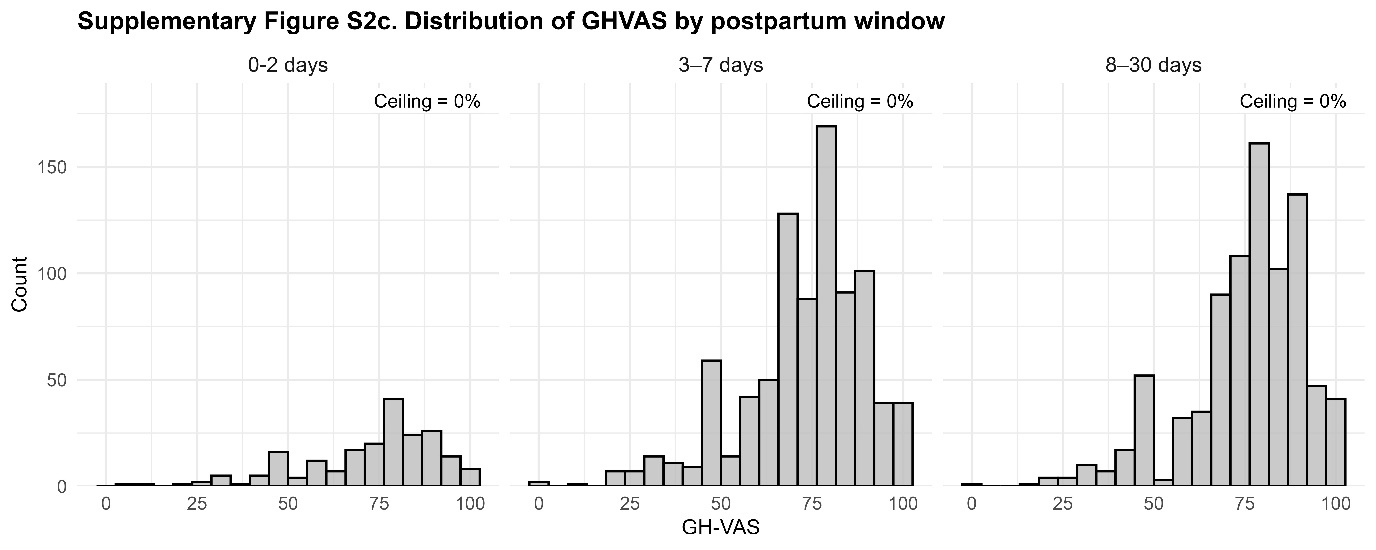

#### **Figure S2d. Histograms of GH-VAS scores by postpartum window, with corresponding ceiling percentage**

*Histograms display the distribution of GH-VAS scores across postpartum windows. Counts are shown on the y-axis. Higher scores indicate better health status.*

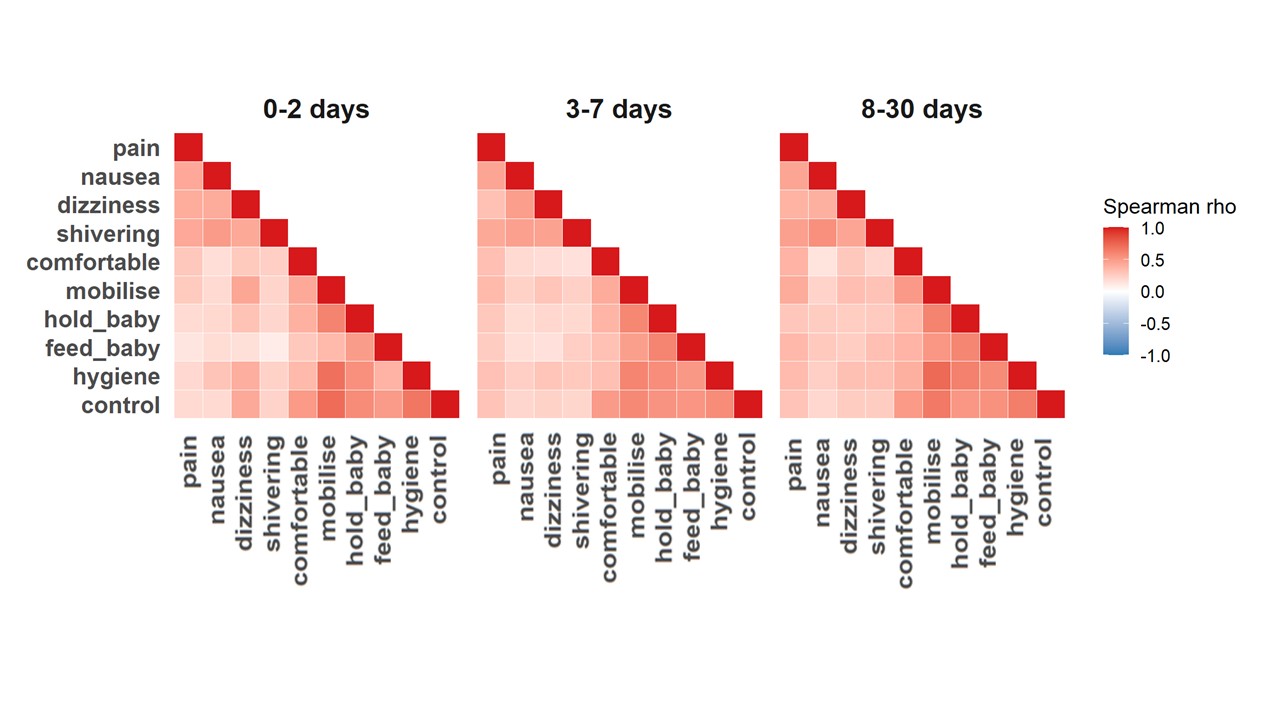

**Figure S2e. ObsQoR-10 item correlations by postpartum windows**

*Heatmaps display Spearman’s rank correlation coefficients (ρ) between ObsQoR-10 items. Colour intensity reflects the magnitude and direction of correlations (scale −1 to 1)*

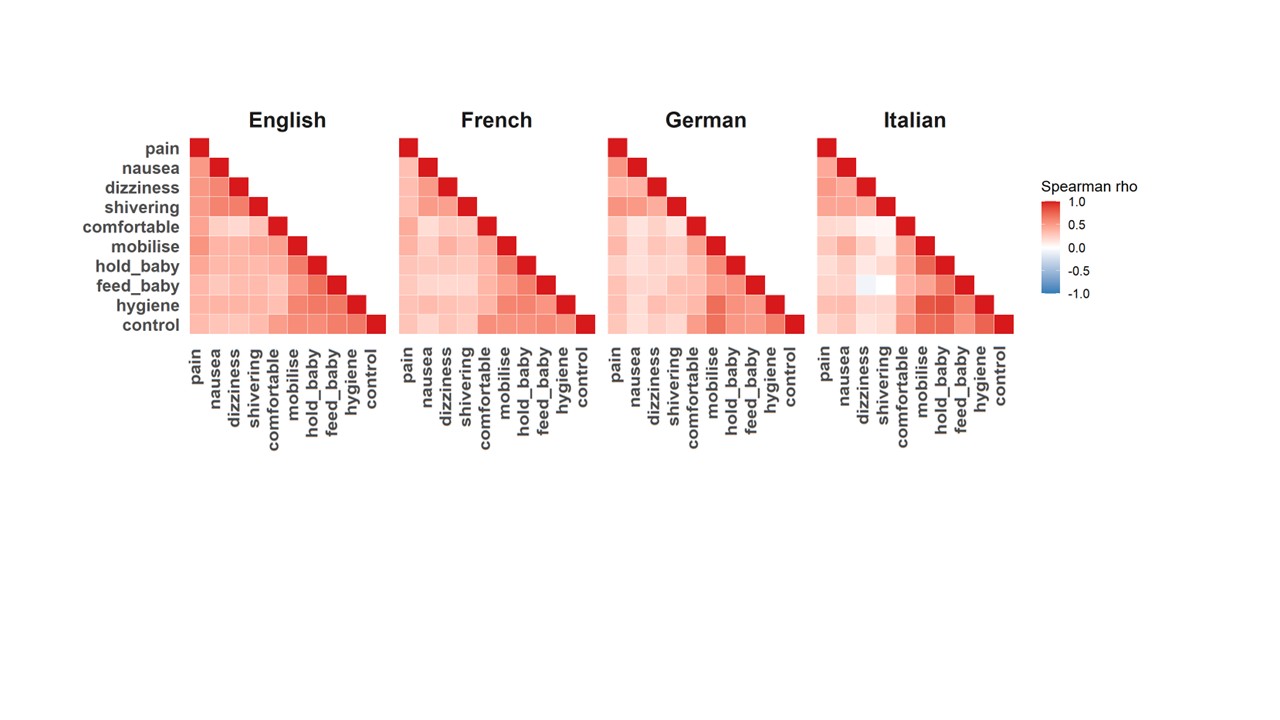

**Figure S2f. ObsQoR-10 item correlations by language**

*Heatmaps display Spearman’s rank correlation coefficients (ρ) between ObsQoR-10 items. Colour intensity reflects the magnitude and direction of correlations (scale −1 to 1)*
